# Vaccination Trends and their Relationship to Continuity of Care in a US-Based Primary Care Practice Registry

**DOI:** 10.1101/2025.09.24.25336567

**Authors:** Nathaniel Hendrix, Marci Nielsen, Andrew Bazemore

## Abstract

Primary care has long played an important role in vaccination in the US and around the world. However, the COVID-19 pandemic disrupted conventional administration patterns for many vaccines, in part through an increased emphasis on mass vaccination sites and pharmacies as a way to expedite the rollout of the COVID vaccine. In an era of increased vaccine hesitancy, though, the frequency of long-term, trusting relationships between patients and their primary care providers may bolster diminishing adherence to recommended vaccine schedules. In this study, we used the American Family Cohort – the US’s largest primary care clinical registry – to assess how patterns of vaccination in primary care have changed since 2017 for ten vaccines recommended on the basis of age. We also assessed how these trends have changed among practices with varying rates of care continuity to test our hypothesis that an ongoing patient-provider relationship improves vaccination rates. In a total of 61,766,185 visits for 6,943,589 patients conducted at 1,323 practices, we observed that the influenza vaccine was the most commonly administered vaccine, followed by pneumococcus. The rates of many vaccines peaked in 2019, though, mirroring national trends that suggest few vaccines are administered at the same rates as they were post-pandemic. We found little support for our hypothesis that care continuity is related to vaccination rates. The population health impact of payment policies and commercial initiatives that shift vaccine administration out of primary care is unclear at this point, but merits ongoing surveillance.

## Introduction

Primary care’s accessibility and broad scope of practice have made it a common platform for the coordination and execution of preventive care, including vaccination. As the count of recommended vaccinations has steadily increased within all phases of patients’ lives,^1^ primary care practice has evolved to address the complexities of modern vaccination schedules. A national survey conducted prior to the COVID-19 pandemic revealed that physicians in primary care provided just under half of the country’s total vaccination-related services – more than any other type of provider or clinical environment.^2^ COVID-19 disrupted the role of primary care clinicians in administering vaccinations, though. In the early phases of COVID-19 vaccination – when speed and scale were the most urgent public priorities – public health departments, hospitals, mass vaccination sites, and pharmacies administered the majority of these new vaccines.^3^

Post-pandemic, vaccination rates continue to decline for children worldwide. In 2023, 2.7 million children were unvaccinated or under-vaccinated, according to the World Health Organization and UNICEF.^4^ In the US, routine vaccination rates for kindergarten children have declined while exemptions from school vaccination requirements, particularly non-medical exemptions, have increased.^5^ This mirrors a global trend of reduced vaccination that has still not recovered to pre-pandemic levels.^6^

The share of missed vaccination attributable to vaccine hesitancy ranges across vaccines from approximately 6% for the birth dose of the hepatitis B vaccine to over 75% for the COVID-19 vaccines.^7,8^ Physicians and other providers in primary care tend to occupy an especially trusted position of information on vaccines and have thus become the subject of special attention as a way to reverse these trends.^9,10^ Information is scarce, though, on the degree to which the COVID-19 pandemic has disrupted the role of primary care providers in administering vaccines. Similarly, quantitative data is lacking on the role of a continuous, trusting relationship with a regular primary care provider in patients’ propensity to receive scheduled vaccinations.

To help answer these questions, we used data from a collection of electronic health records (EHRs) from primary care practices around the United States. Our main objective in this analysis was to assess changes in the annual per-visit probability of receiving one of ten vaccines recommended by the United States Centers for Disease Control and Prevention (CDC) between 2017 and 2023, allowing us to evaluate trends in vaccine administration in primary care before, during, and after the COVID-19 pandemic. As a secondary objective, we explored whether having a regular primary care physician impacts vaccine adherence.

## Methods

### Data Source

We used the American Family Cohort (AFC) for this analysis. The AFC is a research dataset built on the PRIME Registry, which was originally created by the American Board of Family Medicine (ABFM) to provide family physicians and primary care clinicians with efficient tools for practice performance assessment, population health management, risk stratification, and empanelment. Although it is not designed to be representative of all primary care clinicians, it includes records of visits for approximately 8 million unique patients with over 5,000 clinicians, making the AFC the largest clinical registry for primary care in the United States. These records contain comprehensive information on primary health care, including visit patterns, interventions such as medications or procedures, and diagnoses, as well as patient and provider demographics.

Specifically, we used a version of the AFC that had been transformed to the Observational Medical Outcomes Partnership (OMOP) common data model. This harmonized differences in the recording of clinical events that stem either from physician documentation practices or from different EHR platforms. We included visits from January 1, 2017 through December 31, 2023 in this analysis. These dates represent the start of prospective data collection within AFC and the last complete year as of this writing.

### Vaccines

We included ten vaccine types in our analysis: COVID-19; diphtheria, tetanus, and pertussis (DTaP); *Haemophilus influenzae* type B (Hib); hepatitis A; hepatitis B; seasonal influenza; measles, mumps, and rubella (MMR); pneumococcus; varicella; and zoster. We selected these vaccines because each was included in the US CDC’s schedule of vaccines recommended based on age.^11^ Recommendations for some other vaccines such as Mpox or dengue were based on criteria that could not be consistently evaluated in our dataset.

### Outcomes

Our outcome of interest in both analyses was the annual age-adjusted per-visit probability of receiving one of the included vaccines. To arrive at this value, we first identified administrations of each vaccine using procedure codes (Table 1) and text search to identify valid CVX and RxNorm codes associated with vaccines. Once we had the count of vaccine administrations by type, we adjusted the probability of vaccine receipt so that patient age across years (and across continuity subgroups in the secondary analysis) was identically distributed. Specifically, we calculated the sample-wide proportion of visits among patients belonging to 5-year age bins up to a maximum of age 85-plus years old. Then, we standardized the visits of each population to fit the age distribution of visits among the sample-wide population.

**Table 1:**
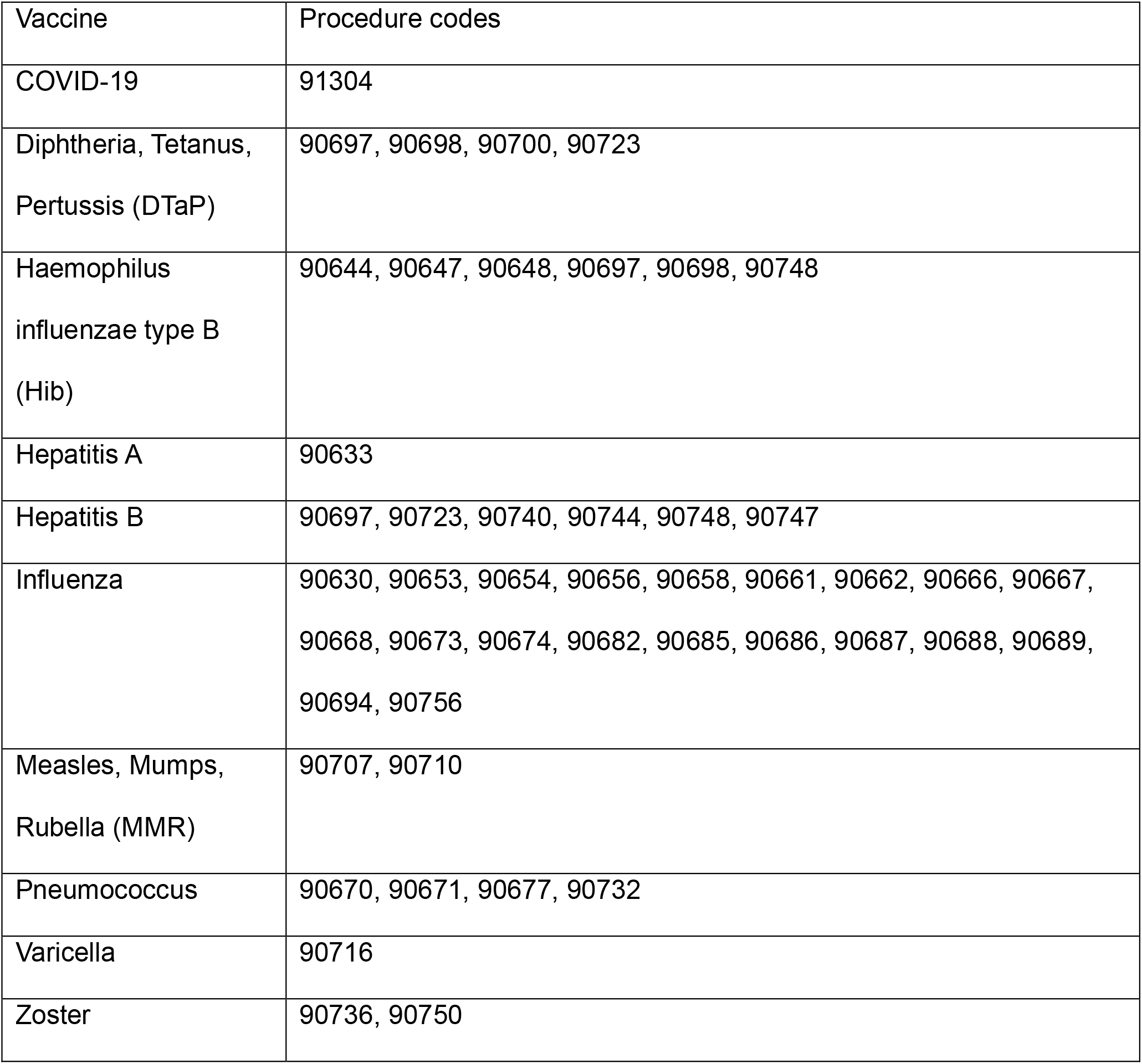
CPT codes for administration of included vaccines.

### Continuity Measure

We used the Bice-Boxerman index^12^ to measure care continuity, which was automatically calculated for each practice in the AFC each year between 2019 and 2021. This index is generally calculated with claims data, where the researcher can observe a patient’s visits to clinicians at different practices. Because we used EHR data and could therefore not reliably observe visits to other clinicians, the measure is constrained to providers within a given practice. This means that it is automatically 100% for each solo practice, which is uninformative. We therefore split the mean continuity score of each practice between 2019 and 2021 into three categories corresponding approximately to tertiles – <80%, 80-99%, and 100% – and added an additional category for solo practices.

### Statistical Analysis

For the analysis of overall rates, we conducted two analyses to identify differences in the annual rate of vaccination within vaccines. First, we used Fisher’s exact test to determine whether vaccination rates vary for a given vaccine across years, as compared to the rate for that vaccine in 2017. Second, we assessed the trend using the Mann-Kendall test, which is a non-parametric measure of a trend’s directionality, strength, and statistical significance relative to no directional trend.

We developed this same strategy further for the analysis of care continuity as a predictor of vaccination rates. We compared the four continuity subgroups first by comparing the mean age-adjusted per-visit probability of vaccination over the entire 2017 to 2023 period, weighted by visit count. For this task, we used ANOVA to determine whether any differences exist between inter-group means and then, if so, used pairwise z-tests to identify which groups were different.

Finally, we sought to identify potential differences in trends between groups by first performing separate Mann-Whitney tests for each category, then employing a standard z-test on Fisher-transformed test results. Given the 54 comparisons made in this analysis – 6 inter-group comparisons within each of the 9 vaccines – we also report Benjamini-Hochberg adjusted p-values.

## Results

Our data contained a total of 61,766,185 visits for 6,943,589 patients from 2017 through 2023. These visits were conducted by 5,001 clinicians at 1,323 practice sites. The most commonly administered vaccine was influenza (Table 2), followed by pneumococcus.

**Table 2:** Raw visit and vaccine count by year.

| Year | Visits | COVID | DTaP | Hib | Hep. A | Hep. B | Influenza | MMR | Pneumo. | Varicella | Zoster |
| --- | --- | --- | --- | --- | --- | --- | --- | --- | --- | --- | --- |
| 2017 | 12,843,213 | 0 | 4,053 | 191 | 996 | 1,351 | 47,888 | 503 | 1,799 | 172 | 0 |
| 2018 | 11,296,632 | 0 | 3,249 | 404 | 1,199 | 1,396 | 52,194 | 422 | 1,929 | 278 | 2,078 |
| 2019 | 10,400,508 | 0 | 3,452 | 368 | 1,265 | 1,517 | 47,220 | 1,367 | 2,080 | 321 | 4,508 |
| 2020 | 8,560,747 | 0 | 2,888 | 246 | 723 | 1,277 | 46,847 | 589 | 1,393 | 260 | 4,460 |
| 2021 | 7,578,269 | 28 | 2,185 | 149 | 310 | 928 | 25,290 | 418 | 782 | 195 | 4,159 |
| 2022 | 6,514,573 | 54 | 1,608 | 81 | 177 | 469 | 14,095 | 196 | 180 | 211 | 698 |
| 2023 | 4,541,396 | 14 | 1,010 | 62 | 98 | 268 | 6,153 | 121 | 134 | 111 | 487 |

Several vaccines such as hepatitis A, MMR, and pneumococcus displayed a notable peak in 2019 (Figure 1). Most differences in rates between years were significant for each of the vaccines. Others, like influenza and varicella, peaked in 2020. Excluding COVID vaccines, only DTaP and varicella vaccines were regularly administered at a significantly higher rate post-pandemic than pre-pandemic. Trends in age-adjusted vaccination rates across this period were non-significant for all vaccines; thus we were unable to reject the null hypothesis of no trend.

**Figure 1:**
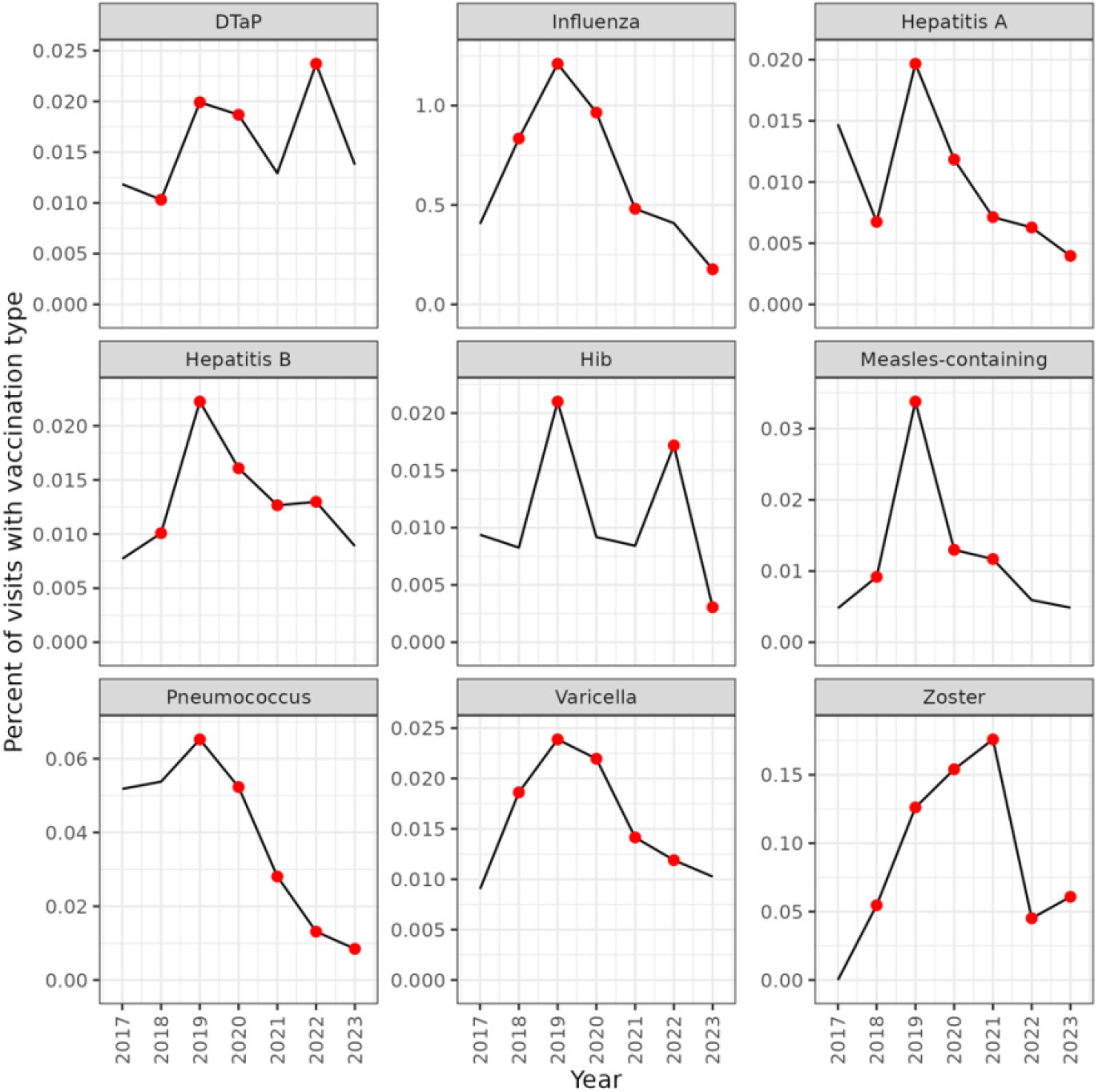
Age-adjusted trends in vaccine administration in primary care practices, with red points indicating years in which the difference from the rate in 2017 was statistically significant per Fisher’s exact test. Note that each vaccine has an independently scaled y-axis and that we did not plot COVID vaccine rates due to very small numbers.

Relatively few practices could be used in the analysis of continuity and vaccine trends due to missing data. Of the 1,323 total practices, 294 (22%) had known practice size of at least 2 clinicians and thus had a calculated continuity score. Practices were nearly equally represented in the three bins of continuity scores: there were 99 practices with a score of 100%; 95 with a score of 80-99%; and 100 with a score less than 80%. An additional 138 were solo practices.

Our a priori hypothesis was that continuity score and vaccine administration would be positively correlated, since the ongoing, trusting relationship between a patient and their primary care provider may help overcome barriers to vaccination. The plots of vaccination rates by continuity score group did not bear this out (Figure 2). Mean vaccination rates were significantly different within the four categories of continuity score for all vaccines except hepatitis A.

**Figure 2:**
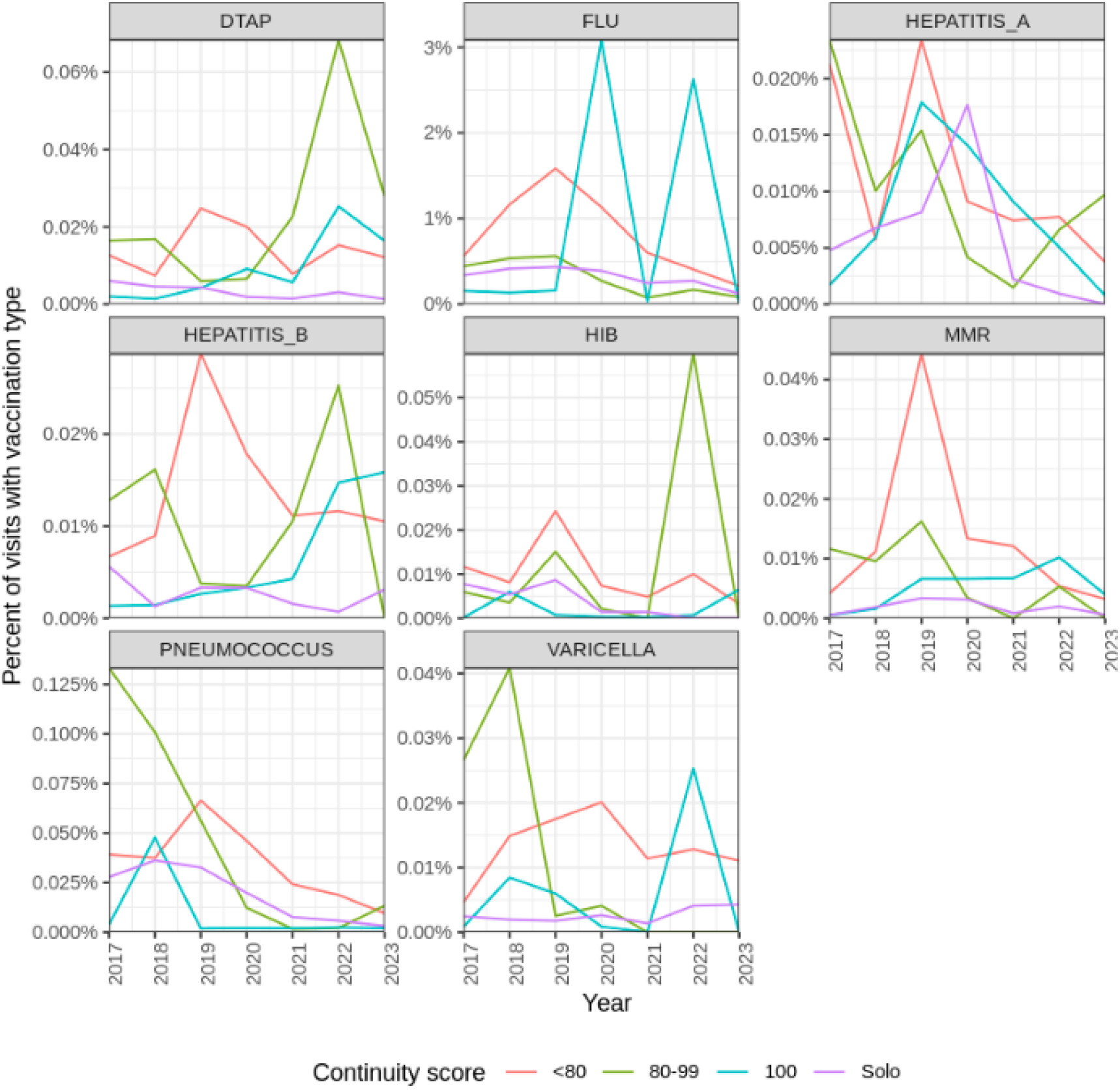
Trends in age-adjusted vaccination probability by binned continuity score. Note that the COVID-19 vaccine was omitted because only solo practitioners administered it.

Among vaccines with significant intergroup differences in age-adjusted mean, most differences were observed between practices in the lowest continuity group and those in either the highest continuity group or solo practices (Figure 3). Those practices in the lowest continuity group, however, often had higher mean vaccination probabilities than other practices. For instance, the observed differences in age-adjusted probability of varicella vaccination suggested a significantly higher rate in practices within the lowest continuity group compared to solo practices and practices in the highest continuity groups, counter to our hypothesis.

**Figure 3:**
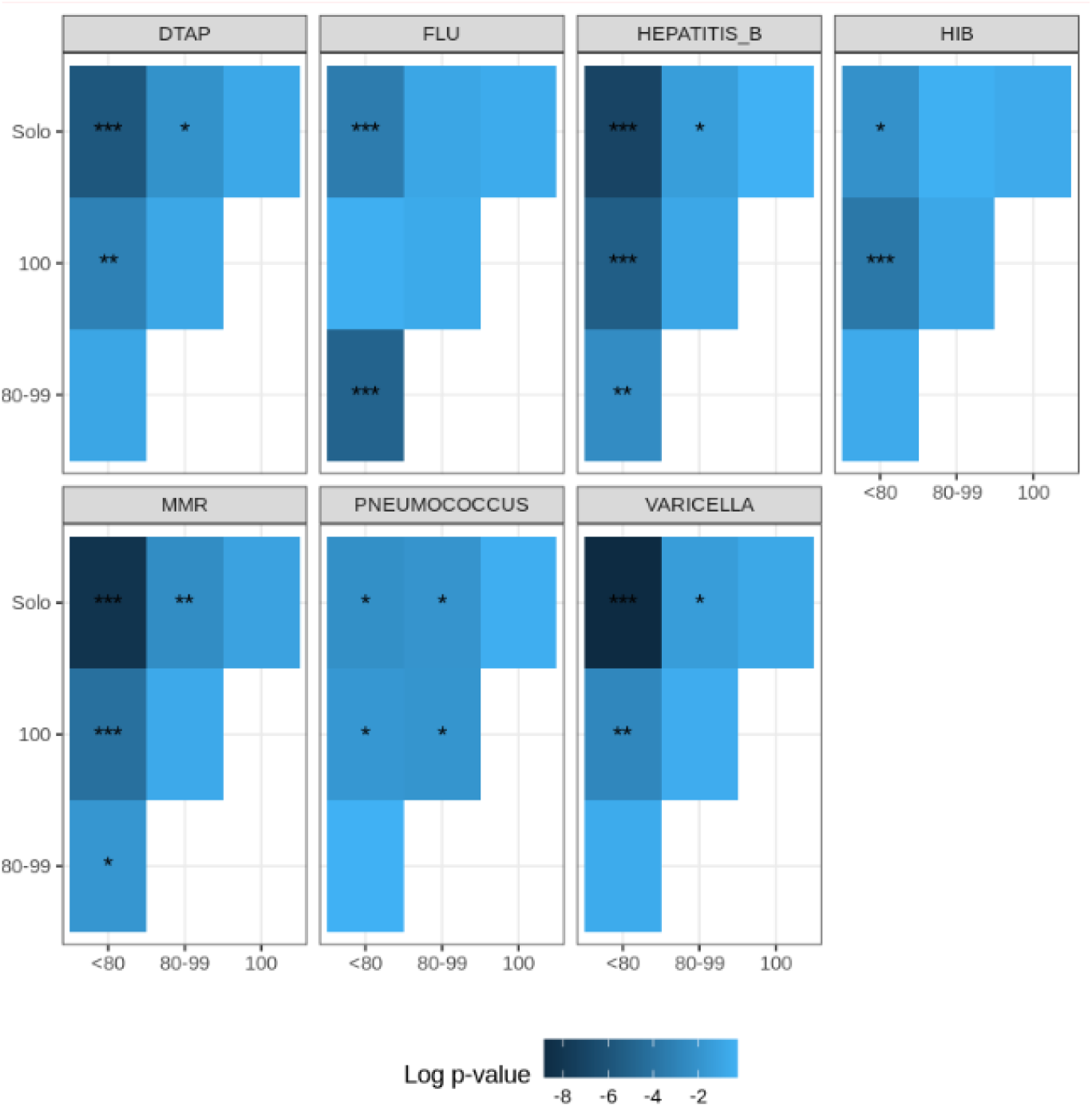
Statistical significance of intergroup differences in mean age-adjusted probability of vaccination (* = p < 0.05, ** = p < 0.005, *** = p < 0.0005).

## Discussion

This novel analysis of vaccination trends from 2017 to 2023 within a US-based national primary care registry reveals several key insights. The decline in vaccination rates post-2020 for most vaccines – except DTaP, Varicella, and HiB – may indicate a significant impact of the COVID-19 pandemic on routine vaccination practices. The most commonly administered vaccine was influenza, which is given annually, followed by pneumococcus, which may align with their prioritization in adult public health campaigns. Notably, vaccination rates for hepatitis A, MMR, and pneumococcus peaked in 2019, while influenza and varicella saw peaks in 2020, potentially reflecting heightened public health initiatives during those periods.

For the most part, our findings directionally mirror national trends in vaccination.^13^ For instance, Hib vaccination has dramatically dropped since 2019. Vaccination rates for varicella, hepatitis B, and MMR have declined more gradually in this same period. Our findings of vaccination rates in primary care do not mirror national trends in vaccination for pneumococcus, though, which has rebounded to pre-pandemic levels nationally; nor do our findings reflect the robust annual increases in zoster vaccination. While pneumococcus is eligible for in-office administration with Medicare Part B, zoster vaccination is not. This observed gap between national trends and vaccination in primary care illustrates how factors such as payment policies and the increased role of commercial pharmacies in vaccination can affect vaccine administration in primary care for a range of vaccines.

The analysis of continuity scores and their association with vaccination trends provided mixed support at best for the hypothesis that higher continuity of care correlates with higher vaccination rates. Practices with higher continuity scores demonstrated more consistent vaccination rates for MMR, hepatitis A, and influenza. Solo practices outperformed other practice types in rates of vaccination against influenza, though underperformed in rates of other vaccines. This provides ambiguous evidence at best for our hypothesis that ongoing, trusting relationships between patients and primary care providers facilitates better adherence to recommended vaccination schedules across the full range of recommended vaccinations. Other studies have found mixed support for this hypothesis as well.^14^

At the same time, we found that lower continuity was associated with higher rates of some vaccinations such as varicella. This finding raises the possibility that the role played by care continuity is contingent on a number of other factors that are likely practice specific. Patient turnover, the prioritization of vaccines, external referral patterns, and local political polarization may all affect how the physician-patient relationship impacts vaccination decisions. Future studies should consider the generalizability of these findings across datasets, as well as the role of socioeconomic factors and urban versus rural practice settings in moderating the effect of care continuity.

The question remains of why patterns were so variable across the different types of vaccines. Most of the vaccines we included in this analysis are administered primarily in childhood, when a substantial share of vaccines are provided to physicians free of charge by the Vaccines for Children program. The Vaccines for Children (VFC) program is a critical public health initiative that provides vaccines at no cost to children who might not otherwise be vaccinated due to their parent’s or guardian’s inability to pay. However, Vaccines for Children has many participation requirements that can be onerous for smaller practices, which leads in turn to high levels of referral to pharmacies and public health clinics among these practices.^15^ Among vaccines administered to older adults, only vaccines against influenza, COVID-19, pneumococcus, and hepatitis B are eligible for in-office administration with Medicare Part B coverage. Additionally, vaccines for COVID-19 have largely been shifted to settings other than primary care clinics.^16^ These limits on the availability and affordability of vaccines administered in primary care settings may incentivize increased use of more convenient settings such as pharmacies. Given the observed heterogeneity in vaccine administration rates, policymakers should consider targeted strategies to support primary care practices, particularly those with lower continuity scores. For example, reducing administrative barriers to the Vaccines for Children program could enable smaller practices to provide broader vaccine coverage. Similarly, expanding Medicare Part B coverage to additional vaccines may improve uptake in adult populations

There were a number of limitations in this study. Our findings about the overall trends in primary care vaccine administration are likely more generalizable than our findings on the impact of care continuity on these trends. We had a relatively small share of practices that had calculable continuity scores, since we relied on EHR rather than claims data. Further research is needed to understand how to recalibrate continuity scores for use with a broader range of practices. It is possible too that the generalizability of our findings is limited by self-selection into our research dataset, which would mean that included practices are not representative of the population of all primary care clinics in the nation. As such, further research should include validation of these trends in other datasets. Finally, the relatively small sample size meant that we could not disambiguate the influence on vaccination trends of payment policies versus pandemic-era disruptions in care provision versus other factors.

This study highlights a general decline in rates of vaccine delivery in primary care practice and considerable heterogeneity across the range of recommended vaccines, along with raising questions about the importance of continuity of care in achieving optimal vaccination coverage. It also reinforces the need for sustained public health efforts to support vaccination in primary care settings.

## Data Availability

Complete data are available from the American Family Cohort for approved research
projects. See up-to-date access requirements at
https://www.AmericanFamilyCohort.org.

## Abbreviations

ABFM: American Board of Family Medicine
AFC: American Family Cohort
CDC: US Centers for Disease Control and Prevention
DTaP: Diphtheria, tetanus, and pertussis
EHR: Electronic Health Record
Hib: *Haemophilus influenzae* type B
MMR: Measles, mumps, and rubella
OMOP: Observational Medical Outcomes Partnership
VFC: Vaccines for Children

